## Supplementary material for "A Rapid Realist Review of the Role of Community Pharmacy in the Public Health Response to COVID-19": Search Terms

### Appendix 1 – Search terms

#### Search log – 28^th^ August 2020

| **Source** | **Date searched** | **Search strategy** | **Hits (or records obtained from searches)** |
| --- | --- | --- | --- |
| **Publish or Perish (Google Scholar)** | 31/07/2020 | Pharmacy[titl] AND Covid* | **715** |
| **Publish or Perish (Google Scholar)** | 31/07/2020 | Pharmacist[titl] AND Covid* | **225** |
| **Ovid MEDLINE** | 31/07/2020 | **exp Coronavirus/ OR exp Coronavirus Infections/ OR (coronavirus* OR 2019‐nCoV OR 2019 ncov OR nCov OR Covid‐19 OR Covid19 OR SARS‐CoV‐2 OR novel coronavirus OR novel corona virus OR covid* or coronavirus 2 OR coronavirus infection* OR coronavirus disease OR corona virus disease OR new coronavirus OR new corona virus OR new coronaviruses OR novel coronaviruses OR wuhan) AND (Community pharmacy OR community pharmacies OR community pharmacist OR community pharmacists OR retail pharmacy OR retail pharmacist OR retail pharmacists OR dispensing chemist OR dispensing chemists OR [(Pharmacist OR pharmacy) AND (community OR “primary care”)] OR dispensary OR apothecary OR druggist)** | **61** |
| **28^th^ August 2020** | EMBASE | 1. **exp Coronavirus/ or exp Coronavirus Infections/ or (coronavirus* or "2019‐nCoV" or " 2019 ncov" or nCov or "Covid‐19" or Covid19 or "SARS‐CoV‐2" or "novel coronavirus" or "novel corona virus" or covid* or "coronavirus 2" or "coronavirus infection*" or "coronavirus disease" or "corona virus disease" or "new coronavirus" or "new corona virus" or "new coronaviruses" or "novel coronaviruses" or wuhan).mp. [mp=title, abstract, heading word, drug trade name, original title, device manufacturer, drug manufacturer, device trade name, keyword, floating subheading word, candidate term word] 78202** 2. **("Community pharmacy" or "community pharmacies" or "community pharmacist" or "community pharmacists" or "retail pharmacy" or "retail pharmacist" or "retail pharmacists" or "dispensing chemist" or "dispensing chemists" or dispensary or apothecary or druggist).mp. [mp=title, abstract, heading word, drug trade name, original title, device manufacturer, drug manufacturer, device trade name, keyword, floating subheading word, candidate term word] 15896** 3. **((Pharmacist or pharmacy or pharmacies) and (community or "primary care")).mp. [mp=title, abstract, heading word, drug trade name, original title, device manufacturer, drug manufacturer, device trade name, keyword, floating subheading word, candidate term word] 26928**   **4. 2 or 3 29215**  **5. 1 and 4 89**  **6. limit 5 to medline 37**  **7. 5 not 6** | **52 hits** |
| **28^th^ August 2020** | CINAHL | **(((( ("Community pharmacy" or "community pharmacies" or "community pharmacist" or "community pharmacists" or "retail pharmacy" or "retail pharmacist" or "retail pharmacists" or "dispensing chemist" or "dispensing chemists" or dispensary or apothecary or druggist) ) OR ( ((Pharmacist or pharmacy or pharmacies) and (community or "primary care")) )) AND exp Coronavirus/ or exp Coronavirus Infections/ or (coronavirus* or "2019‐nCoV" or " 2019 ncov" or nCov or "Covid‐19" or Covid19 or "SARS‐CoV‐2" or "novel coronavirus" or "novel corona virus" or covid* or "coronavirus 2" or "coronavirus infection*" or "coronavirus disease" or "corona virus disease" or "new coronavirus" or "new corona virus" or "new coronaviruses" or "novel coronaviruses" or wuhan)** | **24 hits** |
| **28^th^ August 2020** | Web of Science | **# 4 14 #1 AND #2**  **Refined by: [excluding] Databases: ( MEDLINE )**  **Databases= WOS, BCI, BIOSIS, CCC, DRCI, DIIDW, KJD, MEDLINE, RSCI, SCIELO, ZOOREC Timespan=All years**  **# 3 102 #1 AND #2**  **Databases= WOS, BCI, BIOSIS, CCC, DRCI, DIIDW, KJD, MEDLINE, RSCI, SCIELO, ZOOREC Timespan=All years**  **# 2 32,714 TS=((("Community pharmacy" or "community pharmacies" or "community pharmacist" or "community pharmacists" or "retail pharmacy" or "retail pharmacist" or "retail pharmacists" or "dispensing chemist" or "dispensing chemists" or dispensary or apothecary or druggist) ) OR ( ((Pharmacist or pharmacy or pharmacies) and (community or "primary care") ) ))**  **Databases= WOS, BCI, BIOSIS, CCC, DRCI, DIIDW, KJD, MEDLINE, RSCI, SCIELO, ZOOREC Timespan=All years**  **# 1 95,896 TS=(coronavirus* or "2019‐nCoV" or " 2019 ncov" or nCov or "Covid‐19" or Covid19 or "SARS‐CoV‐2" or "novel coronavirus" or "novel corona virus" or covid* or "coronavirus 2" or "coronavirus infection*" or "coronavirus disease" or "corona virus disease" or "new coronavirus" or "new corona virus" or "new coronaviruses" or "novel coronaviruses" or wuhan)**  **Databases= WOS, BCI, BIOSIS, CCC, DRCI, DIIDW, KJD, MEDLINE, RSCI, SCIELO, ZOOREC Timespan=All years** | **14 hits** |
| **28^th^ August 2020** | Scopus | **( TITLE-ABS-KEY ( coronavirus* OR "2019‐nCoV" OR " 2019 ncov" OR ncov OR "Covid‐19" OR covid19 OR "SARS‐CoV‐2" OR "novel coronavirus" OR "novel corona virus" OR covid* OR "coronavirus 2" OR "coronavirus infection*" OR "coronavirus disease" ) ) AND ( ( ( ( "Community pharmacy" OR "community pharmacies" OR "community pharmacist" OR "community pharmacists" OR "retail pharmacy" OR "retail pharmacist" OR "retail pharmacists" OR "dispensing chemist" OR "dispensing chemists" OR dispensary OR apothecary OR druggist ) ) OR ( ( ( pharmacist OR pharmacy OR pharmacies ) AND ( community OR "primary care" ) ) ) ) )** | **591 hits** |
| **28^th^ August 2020** |  | **Search of the contents of preprint services and the WHO Covid register: pharmacy OR pharmacist OR pharmacists or pharmacies** | **(402 hits)** |
| **28th August 2020** |  | **e.g. the curated BioRxiv/MedRxiv dataset (connect.medrxiv.org/relate/content/181).** | **10 hits and 19 hits – none relevant** |
| **28th August 2020** |  | **Social media (e.g. blogs) including facilitated twitter® discussion (#Cpharmchat) on the Impact of COVID-19 on community pharmacy.** | **Twitter 8 hits** |
| **28th August 2020** |  | **Web-sites/emails from relevant regulators and professional organisations including RPS (https://www.rpharms.com/), PDA (https://www.the-pda.org/), PSNC (http://psnc.org.uk/), and GPhC (https://www.pharmacyregulation.org/).** | **Completed** |
| **28th August 2020** |  | **Pharmaceutical Journal web-site** [**https://www.pharmaceutical-journal.com/**](https://www.pharmaceutical-journal.com/)**. No language restrictions at title and abstract stage selection stage. Separated high and low/middle income countries at sift stage.** | **Completed** |
| **28th August 2020** |  | **Local Pharmaceutical Committees (LPC)** | **Completed** |
