## Supplementary material for "A Rapid Realist Review of the Role of Community Pharmacy in the Public Health Response to COVID-19": Literature inclusion criteria

Appendix 2 - Literature inclusion criteria

Published after January 2003 AND

COVID-19 or other pandemic or other infectious diseases

OR vaccination programmes

OR expanded/extended roles

AND Community Pharmacy

AND High- or middle-income country
