## Supplementary material for "A Rapid Realist Review of the Role of Community Pharmacy in the Public Health Response to COVID-19": PRISMA 2009 Flow Diagram

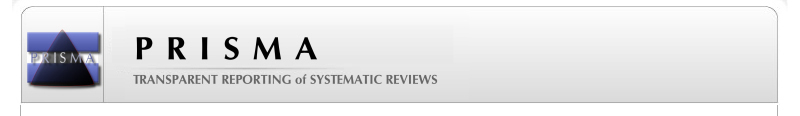
**PRISMA 2009 Flow Diagram**

**Screening**

**Included**

**Eligibility**

**Identification**

Records identified through database searching
n = 2083

Additional records identified through other sources
(n = 78)

Records after duplicates and older records removed (n = 1532)

Records screened on title and abstract
(n = 1532)

Records excluded
(n = 1300)

Not COVID; not other pandemic or other infectious diseases; not community pharmacy related; low or middle income country

Full-text articles assessed for eligibility
n = 232

Full-text articles excluded, with reasons
n = 129

Did not contribute to the development or refinement of CMOCs

Studies included in qualitative synthesis

n = 103
