## Supplementary material for "A Rapid Realist Review of the Role of Community Pharmacy in the Public Health Response to COVID-19": Programme Theory

Appendix 4 - A simplified diagram of the programme theory of a COVID-19 vaccination programme provided by community pharmacies

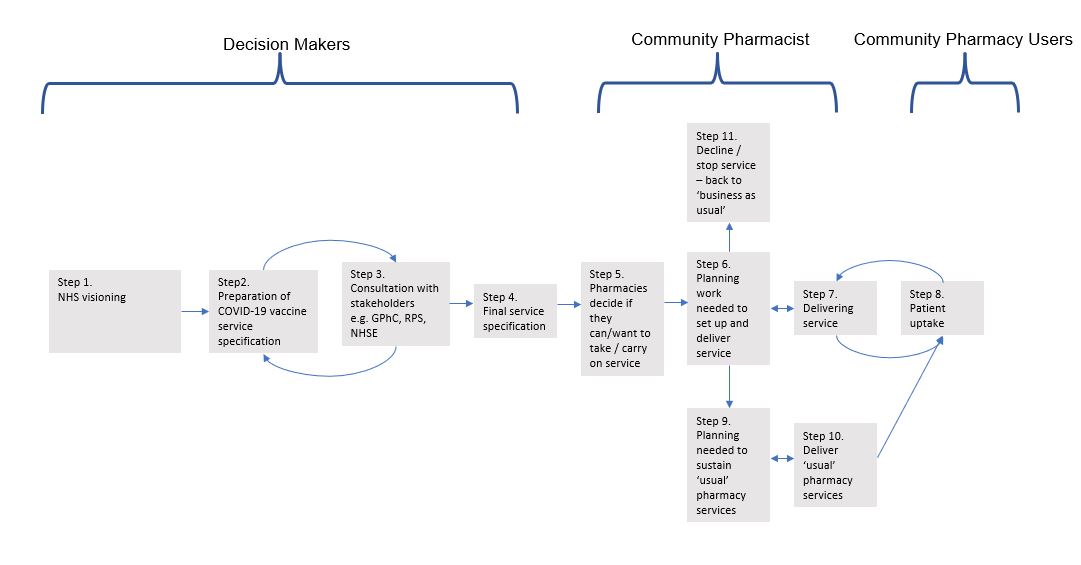
